# Targeted memory reactivation during REM sleep in patients with social anxiety disorder

**DOI:** 10.1101/2022.03.19.22272647

**Authors:** F. Borghese, P. Henckaerts, F. Guy, C. Mayo, S. Delplanque, S. Schwartz, L. Perogamvros

## Abstract

**Background:** Social anxiety disorder (SAD) is an anxiety disorder characterized by a significant amount of fear when confronted to social situations and can cause considerable distress in daily life. Exposure therapy, which is based on fear extinction, is a popular and effective treatment for SAD, although it does not often lead to full remission. Here, we aimed at improving exposure therapy outcome. Specifically, based on previous research showing that rapid eye movement (REM) sleep promotes the consolidation of extinction memory, we used targeted memory reactivation (TMR) during REM sleep to enhance extinction learning.

**Methods:** 48 subjects (32 women and 16 men, mean age of 24.41 ± 4.91) with moderate or severe SAD according to DSM-5 participated in our study, and were randomly assigned to one of two matched groups: control or TMR group. All patients had two successive exposure therapy sessions in a virtual reality (VR) environment, where they were asked to give a public talk in front of a virtual jury. At the end of each session, and only in the TMR group (N=24), a sound was paired to the positive feedback phase of exposure therapy (i.e. approval of their performance), and which represents the extinction memory to be strengthened during REM sleep. All participants slept at home with a wearable headband device which automatically identified sleep stages online and administered the sound several times during REM sleep. Anxiety level was assessed using measures of sympathetic (electrodermal activity component : non-specific skin conductance responses, ns-SCRs) and parasympathetic (heart rate variability component : root mean square of successive differences between normal heartbeats, RMSSD) activity, and subjective measures (Subjective Units of Distress Scale, SUDS), during the preparation phase of their virtual talks before (T1) and after (T2) one full-night’s sleep with auditory stimulation and after one week of auditory stimulation at home (T3). Participants also filled in a dream diary one week prior and one week after the day of exposure therapy.

**Results:** Subjective anxiety was reduced during the second and third anticipatory preparation phase of exposure, compared to the first one, for both groups (p < 0.001). RMSSD levels were lower in the TMR group compared to the control group (p=.037) during the preparation phase after 8 nights of stimulation at home (T3). No significant result between groups was observed for SUDS and the ns-SCRs at T3. Importantly, the longer REM sleep and the more stimulations the TMR group (but not the control group) had at home, the less anxious (increased RMSSD) these participants were. Finally, fear in dreams correlated positively with measures of stress (ns-SCRs and SUDS) in this group.

**Conclusions:** TMR during REM sleep did not modulate the beneficial effect of exposure therapy on anxiety-related distress (SUDS). Yet, our results support that REM sleep can contribute to extinction processes and substantiate strong links between emotional experiences in dreams and waking stress levels in these patients.

## 1. Introduction

Social anxiety disorder (SAD) is characterized by an exaggerated and persistent amount of fear when confronted with social situations (Leichsenring & Leweke, 2017), which can lead individuals with SAD to either avoid such situations or endure them with significant discomfort (Leichsenring & Leweke, 2017; Stein & Stein, 2008). It is a chronic disorder with a lifetime prevalence rate of 13% and an early onset in adolescence (Bandelow & Michaelis, 2015; Morrison & Heimberg, 2013). Studies looking into the mechanisms underlying anxiety disorders converge to suggest that these disorders are characterized by dysfunctional fear extinction (Craske et al., 2018).

Fear conditioning is a form of associative learning between a neutral stimulus (conditioned stimulus CS) and an innately aversive stimulus (unconditioned stimulus US), after repeated pairings of the CS and the US. After fear conditioning, presentation of the CS alone triggers an emotional response (conditioned response, CR) (Graham & Milad, 2011; Neumann & Waters, 2006). Fear extinction learning (or inhibitory learning) is a process during which the conditioned fear response decreases or is inhibited when there is a repeated presentation of the CS in the absence of the US. Existing data support that extinction induces the learning of a new association ‘CS-noUS’, in which the CS no longer predicts the US (Bouton, 2002; Craske et al., 2018). Inhibitory learning is central to extinction and deficit in this process could contribute to the development of anxiety disorders (Craske et al., 2014; Graham & Milad, 2011).

Exposure therapy is a treatment based on extinction learning mechanisms and involves the gradual and repeated exposure to the feared stimuli in the absence of the negative outcome (Craske et al., 2018). It is a popular and efficient treatment of SAD and other anxiety disorders such as generalized anxiety disorder (GAD) and specific phobias (Kaczkurkin & Foa, 2015). However, while there is a consensus in the field of psychotherapy regarding the efficiency of exposure therapy to treat SAD, patients are reluctant to seek treatment, as they can be hesitant to engage in social interactions. The use of virtual reality (VR) allows a better control of exposure conditions and the ability to stop or take breaks if the patient is overwhelmed (Bouchard et al., 2017). A meta-analysis conducted by Carl et al. (2019) indicated that virtual exposure and in vivo exposure showed similar efficacy in the treatment of anxiety disorders (including SAD), both leading to reduction of anxiety symptoms. However, exposure is not a foolproof solution to treat anxiety disorders, as its efficacy is not always significant in the long run. Many patients experience a return of fear at the end of the exposure therapy, with rates up to 62% (Craske et al., 2018). Therefore, there is an emerging need to find ways to enhance the therapeutic outcome of this therapy.

In order to enhance extinction learning, before, during or after exposure therapy, several methods have been used, such as administration of cortisol (Soravia et al., 2014) or stimulation of medial prefrontal cortex (mPFC) with repetitive transcranial magnetic stimulation (Raij et al., 2018). A simple positive reinforcement or feedback (e.g. positive compliments) regarding the patient’s performance, which represents extinction-related violation of expectancy (Craske et al., 2018), was also found to reduce social anxiety (Hartanto et al., 2014), and is an integral part of self-focused exposure therapy (Borgeat et al., 2009). Other protocols have used periods of sleep (naps or full night’s sleep) to reinforce the consolidation of extinction learning after exposure therapy for anxiety disorders, such as spider phobia (Kleim et al., 2014; Pace-Schott et al., 2012). A preliminary study recently demonstrated that naps after exposure therapy for SAD lowered sympathetic responses (as measured by skin conductance response and cortisol levels) during anticipation of a social challenge at a trend level (Pace-Schott et al., 2018). In their conclusion, these authors suggested that the lack of significance could potentially be due to the fact that sleep post-exposure was not long enough for the participants to experience rapid eye movement (REM) sleep, and that REM sleep could play an important role in the consolidation of extinction learning.

Indeed, accumulating evidence shows that REM sleep may represent a permissive condition for the processing of extinction memory. Healthy participants who had REM sleep after extinction learning exhibited greater extinction recall, accompanied by stronger activation of the ventromedial prefrontal cortex (vmPFC) in response to the extinguished stimulus (Spoormaker et al., 2012; Spoormaker et al., 2010). Moreover, a greater retention of extinction learning was associated with REM percent in an intervening overnight sleep (Pace-Schott et al., 2014), while a subsequent study elegantly demonstrated that REM sleep (but not slow wave sleep-SWS or wakefulness) causes successful consolidation of extinction memory (Menz et al., 2016). Other studies also showed that REM sleep helps to decrease the experienced arousal or affective tonus associated with emotional events, thus leading to higher familiarity and habituation to emotionally negative stimuli (emotional depotentiation) (Gujar et al., 2011; van der Helm et al., 2011). Recent neuroimaging data also indicated that negative emotions in dreams, and specifically fear, may contribute to (or reflect) emotional regulation processes during sleep and yield better adapted responses to aversive stimuli during waking life (Sterpenich et al., 2020).

Memory reactivation during sleep can be induced or intensified with targeted memory reactivation (TMR). This method consists in associating a sensory cue with a learning experience, and subsequently presenting this cue to increase the likelihood that the memory of this experience is reactivated. Thus, presenting the cue during sleep will trigger a neuronal replay of the associated memory, which will strengthen memory consolidation (Oudiette & Paller, 2013). Studies have demonstrated that such cued memory reactivation can improve the consolidation of declarative and procedural memories to levels up to 35% as compared to wakefulness (Diekelmann, 2014; Klinzing & Diekelmann, 2019; Schouten et al., 2017). While the benefits of applying TMR during non-REM (NREM) sleep have been established across many studies, results are more inconsistent when cueing occurs during REM sleep (Schouten et al., 2017). TMR during REM sleep enhanced memory (Guerrien et al., 1989; Smith & Weeden, 1990), including associative memory (Sterpenich et al., 2014)(contrary to SWS ; Ashton et al. (2018)), and increased positive valence of negative stimuli (Rihm & Rasch, 2015; Walker & van der Helm, 2009).

The main goal of this study was to investigate whether TMR during REM sleep may enhance exposure therapy in SAD. Even though REM sleep and dreaming appear to have an important role in extinction learning and emotional depotentiation, to our knowledge, no study to date used TMR during REM sleep in the context of treating anxiety disorders. In our study, participants with SAD took part in virtual reality exposure sessions during which they were asked to perform public presentations.

Anxiety levels were assessed at different time points with subjective and physiological measures. Specifically, the anticipatory phase of their performance being particularly stressful (Carrillo et al., 2001; Gonzalez-Bono et al., 2002; Tardy & Allen, 1998), was chosen as a critical period for stress measurement. Following the public presentations in VR, participants received positive feedback of their performance (Borgeat et al., 2009), which served as an extinction period of the therapy. Indeed, as there are no negative consequences such as negative judgment from the jury following the feared situation, this period represents a CS-noUS association reminiscent of extinction learning (Craske et al., 2018). During this period, participants in the experimental group (TMR group) were exposed to an auditory cue, while those in the control group were not. In the frame of the TMR technique, this allows for an association between the sound and the extinction memory. During eight nights following the first virtual exposure session, participants from both groups received the auditory cue selectively during REM sleep.

The primary hypothesis of this study was that (1) participants in the TMR group will show reduced intensity of social anxiety compared to participants in the control group, based on subjective reports and physiological measures, after eight nights of stimulations during REM sleep. Secondary hypotheses included the following: (2) the aforementioned effect could be already observable after one night of sound presentation during REM sleep; (3) participants in the TMR group will experience generalization of extinction compared to participants in the control group, after eight nights of sound presentation during REM sleep; and (4) the number of stimulations in the TMR group and/or REM sleep duration will have a beneficial effect on stress, respectively due to increased TMR-related events and the proposed role of REM sleep in extinction learning and emotional depotentiation. Considering recent evidence on the links between dreams and emotional processing in wakefulness (Sterpenich et al., 2020), we also hypothesized that (5) fear in dreams should positively correlate with reduced stress in wakefulness.

## 2. Methods

### 2.1. Participants

Participants were recruited through flyers and advertisements on social media. Inclusion criteria were: being aged between 16 and 40 years old, having SAD according to the Diagnostic and Statistical Manual of Mental Disorder, 5th edition (American Psychiatric Association, 2013), not being under treatment for social anxiety, and not presenting other mental disorders or sleep disorders. Patients with symptoms of obstructive sleep apnea syndrome, restless legs syndrome, insomnia disorder, or under anxiolytics, antipsychotic or antidepressant medication were excluded. Initially, 51 participants were recruited. Three of them were not included in the final analyses because they withdrew from the study. The final sample of participants was composed of 48 participants (32 females and 16 males) with SAD, as assessed by an interview with a certified psychologist. Participants gave their written consent to take part in this study and received a participant fee. The study was approved by the ethical committee of the canton of Geneva, Switzerland (*‘Commission Cantonale d’Ethique de la Recherche sur l’être humain’*).

### 2.2. Procedure

We used a single-blind randomized controlled design, with the assignment of 48 SAD patients into two groups (TMR group and control group, Fig. 1). This randomization took place on Day 1 (T1). Patients of the TMR group received a neutral auditory stimulus (i.e., a 1-second piano chord (C69)), which was associated with the positive feedback phase of exposure therapy. The aim was to consolidate this new associative memory during REM sleep through TMR. After this session, all participants had a full night of polysomnography (PSG). During sleep, patients of both groups received the sound during REM sleep with a wireless sleep headband. On Day 2, the participants underwent another VR session of exposure therapy (T2). Then, they spent one week at home with the headband device administrating the sound during REM sleep. On day 9, participants came for one last VR session of exposure therapy (T3). Social anxiety for a public talk was measured during the preparation of the talk at six different time points: before (T1a) and after (T1b) the first session of VR exposure therapy, after sleep with TMR (T2), and after 1 week of TMR at home (T3). In order to test for generalization of extinction, we tested anxiety for another context (being approached by virtual characters) at time points T0 and T4. Throughout all phases of the VR sessions (baseline, preparation, presentation, positive feedback), anxiety was measured at the subjective level (Subjective Units of Distress Scale) and physiological level, including heart rate variability and electrodermal activity.

**Figure 1.**
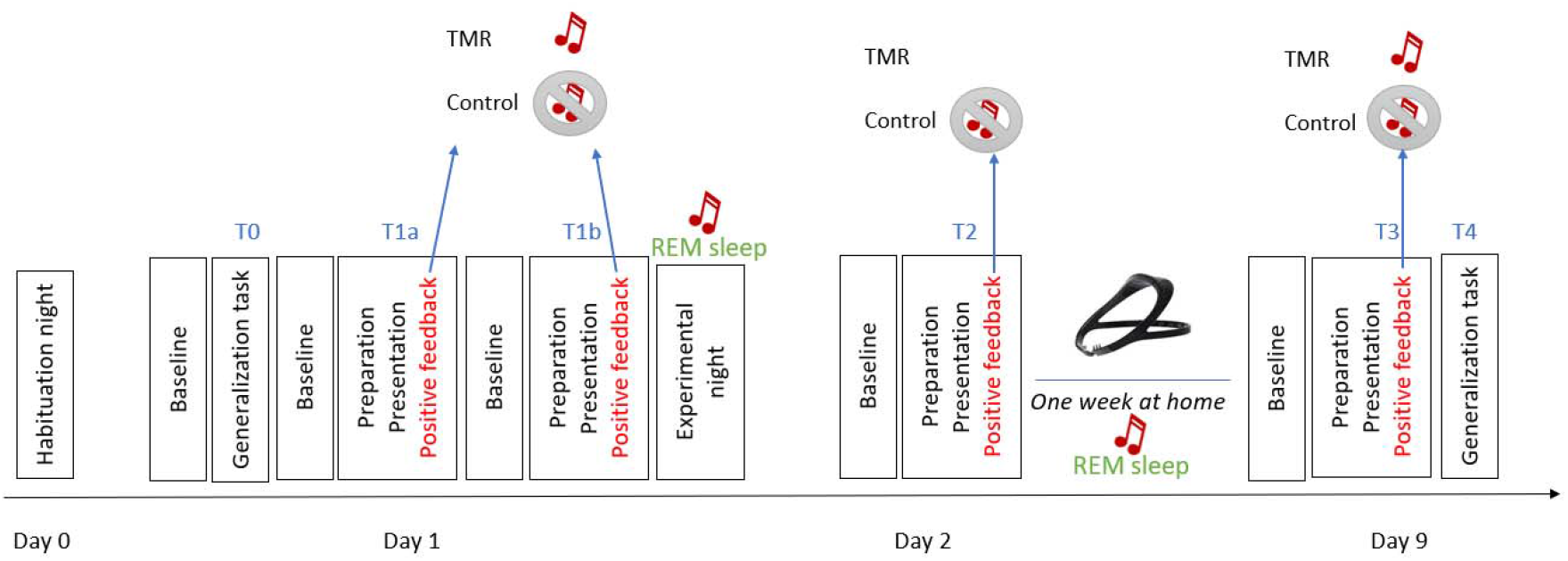
Study design. Participants underwent a habituation night on Day 0. The following day (Day 1), they had one VR session of generalization task (T0) and two VR sessions of self-focused exposure therapy (T1a and T1b), before the experimental night. On Day 2, participants underwent another VR session of self-focused exposure therapy (T2). Then, they spent one week at home with a headband sleep device. On day 9, participants came for one last VR session of self-focused exposure therapy (T3) and one VR session of generalization task (T4). During the positive feedback phases of the VR, a sound was administered to the TMR group, while no sound was administered to the control group. Both groups were administered the sound during their REM sleep at the experimental night and during the nights at home with a sleep headband.

### 2.3. Materials

#### VR environment

Participants were immersed in a virtual environment, with an Oculus Rift Headset (Meta Quest, Irvine, California, United States). The virtual environment was designed on Unity 3D (Unity Software Inc., San Francisco, United States) by the Virtual Reality & Robotics facility of the Human Neuroscience Platform, Fondation Campus Biotech Geneva. The virtual environment consisted of a theatre stage and an audience space. On the stage, there was a microphone stand in front of which the participant made his/her presentations. From the stage, the participants could see the audience space, where there was a table (with a timer on it), two jury members (one male and one female) and an audience of about twenty members. Electrodes for the recording of electrodermal activity and electrocardiogram signals were placed on the participant before he/she entered the booth. The participant could communicate and hear the instructions from the experimenter through the headphones and microphone included in the VR headset.

#### Structure of each VR exposure session

All the VR sessions of the main task (T1a, T1b, T2, T3) in this experiment have the same structure (see Supplementary Material S2 for detailed description). They started with a three-minute baseline phase during which participants had to stay seated, relaxed and get used to the environment (**baseline phase**). The goal of this rest phase was to allow any distress the participant might feel to dissipate. After this phase, they were allowed 5 minutes to mentally prepare a short speech on a given topic (**preparation phase**), which they would present afterwards for 5 minutes in a theater room with a two-person jury and a public in the background (**presentation phase**). At the end of their performance, a virtual feedback was given to them: first, a standard positive feedback from the virtual jury (1.5 min), and then the experimenter gave an individualized positive feedback (3.5 min). During this phase (**positive feedback phase**), a sound was administered every ten seconds in the TMR group, while no such sound was administered in the control group.

#### Sleep headband Dreem®

The Dreem® headband is a wireless sleep headband designed by the company Dreem (Dreem SAS, Paris, https://dreem.com/en/product). It is composed of fabric and foam and can be adjusted with an elastic band behind the head. The device records and analyses physiological data: (1) brain activity via EEG dry electrodes (derivations: FpZ-O1, FpZ-O2, FpZ-F7, F8-F7, F7-O1, F8-O2, FpZ-F8; 250 Hz with a 0.4–35 Hz bandpass filter), (2) movements, (3) sleep position, and (4) heart rate. The EEG electrodes are placed at the front and back of the device. Sound can be delivered via bone-conduction transducers integrated in the headband. The headband monitors sleep and can detect different sleep stages through a reliable algorithm (Arnal et al., 2020). In our experiment, whenever REM sleep was detected for more than five minutes, the sound was delivered to the participants every 10 seconds. These auditory stimulations were interrupted whenever a new sleep stage was detected (after which they restarted after 5 minutes of REM), after detection of a movement (90 seconds of interruption), after detection of an alpha wave (45 seconds of interruption), after detection of a blink (10 seconds of interruption), and after bad quality signal (4 seconds of interruption). The headband was connected to a smartphone application, which provided a summary on sleep quality, and allowed each participant to choose the volume of the sound.

#### Questionnaires

As part of the recruitment, participants filled several online questionnaires used to verify inclusion criteria: general socio-demographic and medical questionnaires, the Liebowitz Social Anxiety Scale (LSAS) (Liebowitz, 1987), the Pittsburgh Sleep Quality Index (PSQI) (Buysse et al., 1989), the Beck Anxiety Inventory (BAI) (Beck et al., 1988), the Beck Depression Inventory II (BDI II) (Beck et al., 1996), the Insomnia Severity Index (ISI) (Morin et al., 2011). During initial assessment, a certified psychologist used the Mini-International Neuropsychiatric Interview (MINI) module on social anxiety disorder (Sheehan, 2015), a short structured diagnostic interview instrument, based on the DSM-5 criteria.

#### Actigraphy, dream diary, sleep agenda

During two weeks (starting from one week before the first VR session and finishing at the end of the protocol), participants wore an actigraph, and filled out a dream diary and a sleep agenda. Among other information, the dream diary contained some specific questions concerning which emotions were present in the participants’ dreams (see also Sterpenich et al. (2020)). Before each VR session, participants also performed a psychomotor vigilance task (PVT). After the two nights at the lab, and after one week of sleeping at home, participants filled out post-sleep the St. Mary’s Hospital questionnaire (Ellis et al., 1981) to evaluate the quality of their sleep.

### 2.4. Measurements

#### Measures of anxiety

During the VR tasks, we assessed stress levels with two physiological measures: heart rate variability (HRV) (Alvares et al., 2013; Pulopulos et al., 2018; Wendt et al., 2015) and electrodermal activity (EDA) (Boucsein et al., 2012; Wilhelm et al., 2005a), which provide estimates of parasympathetic and sympathetic nervous system activity respectively. EDA was measured with disposable adhesive sensors on the distal phalanges of the index and middle finger of the non-dominant hand of participants. EDA was analyzed to obtain skin conductance responses (SCRs). For our study, we were interested in non-specific SCRs (ns-SCRs), which were counted during each VR phase, including the preparation period of the talks (target phase for the physiological outcome measures). HRV was measured using three ECG electrodes (including one ground) placed below the rib (left), under the clavicle (right) and on the hip (right) for the ground. For our study, we were interested in the root mean square of successive differences between normal heartbeats (RMSSD, in ms) for each VR session phase. The RMSSD is used to estimate the vagally mediated changes in heart rate variability (HRV) (Shaffer & Ginsberg, 2017) and presents the advantage of being relatively free of respiratory influences compared to other variables calculated from HRV (Laborde et al., 2017). Both ns-SCRs and RMSSD were recorded measured using the Biopac MP160 System, (Biopac Systems Inc., Goleta, CA, 2013) and the software AcqKnowledge v.5.0. Finally, the adapted Subjective Units of Distress Scale, SUDS (Wolpe, 1969, 1990) was used during the Virtual Reality (VR) sessions to measure the intensity of distress of people suffering from social anxiety. It consists in a single question scale, in which the subject rates on a scale from 0 to 10 the level of distress that they were feeling at a specific moment. The SUDS was given after each positive feedback and preparation phase, after T0, as well as before the preparations T2 and T3. Details of data analysis of ns-SCRs and RMSSD are provided in Supplementary Material (S1).

#### Polysomnography

To assess the sleep structure of the participants we recorded polysomnography (PSG) for two nights at the sleep laboratory (one habituation night without auditory stimulations, one experimental night with auditory stimulations). There were six electrodes (F3, F4, C3, C4, O1 and O2) placed on the head using the 10-20 system. We also put two electrodes to measure the eye movements (EOG1 and EOG2) and three electrodes to measure muscle tone (EMG1, EMG2 and EMG3). We placed the references on the mastoids and the ground was placed on the cheekbone. We also placed two electrodes to measure the heart rate (ECG). The setup and sleep scoring was done according to the AASM manual guidelines (Berry et al., 2012).

#### Primary and secondary outcome measures

The primary psychophysiological outcome measure was the RMSSD, during the preparation phase of exposure therapy (Laborde et al., 2017). Lower levels of RMSSD indicate higher anxiety (Francis et al., 2009), however, we need to keep in mind that RMSSD levels can be modulated by other factors than stress or anxiety, such as physical activity, depression, worry and measures of inflammation (McCraty & Shaffer, 2015). The primary subjective measure of distress was the SUDS collected immediately after the end of the preparation phase and before the oral presentation in the VR environment.

The secondary outcome measure was the ns-SCRs (Carrillo et al., 2001) during the preparation phase of exposure therapy, used here as a physiological indicator of anticipatory anxiety (Boucsein et al., 2012). Other exploratory variables included the change of fear in dreams (average of fear during the second week with stimulations minus the first week without stimulations).

The anticipatory phase of social performance (e.g. preparation of public speaking) was chosen as the main phase to study social anxiety, as it has been shown to be particularly stressful (Cornwell et al., 2006).

#### Sample size consideration

Based on a previous study (Gaebler et al., 2013) on the difference of SAD vs control group for HRV with an effect size of d=0.77, 44 patients (22 per arm) would be required to have an 80% chance of detecting this difference between patients with TMR vs. those in the control group. Besides, based on a study (Sterpenich et al., 2014) on the effect of associated vs non-associated sound during REM sleep on associative memory with a large effect size (g=1.01), this sample size would be sufficient for the aforementioned detection.

### 2.5. Statistical analysis

#### Levels of anxiety

These analyses were done on 46 participants (2 participants from the initial sample were excluded due to unusable data). Data were entered into a multilevel regression model, with either the number of RMSSD levels or SUDS scores, as dependent variables, and time and group as (interacting) independent variables. The latter represented the fixed effects of the multilevel model, while random effects were represented by a random intercept for subjects (`Y ∼ group*time + (1|ID)`). The random intercept accounted for correlation between repeated measures, by assuming baseline differences between subjects in the average DV. A multilevel regression was chosen for these data, due to its ability to handle (a) missing data in the time variable, (b) time-varying covariates, and (c) continuous within-subject covariates, and (d) continuous within-subject covariates. Once the model was fitted, we performed a Type II ANOVA breakdown of fixed effects using *F*-tests, starting with the interaction test of time × group, followed by main effects tests for time and group separately. Significant effects were further explored with pairwise *t*-tests. Degrees of freedom for all *F*- and *t*-tests were adjusted for the random effects structure using Satterthwaite’s method (Fitzmaurice et al., 2004) yielding fractional degrees of freedom. Multilevel analyses were conducted with the R statistical language, version 1.2.5019, (RStudio Team, Boston, MA, 2019), using the packages “lme4” for model estimation (Bates, Machler, Bolker, & Walker, 2015) and “lmerTest” for inferential tests (Kuznetsova, Brockhoff, & Christensen, 2016). The secondary psychophysiological outcome measure, i.e. ns-SCRs, underwent the same analysis structure.

#### Sleep variables

Data on several sleep variables (e.g., duration of sleep stages, total sleep time, number of auditory stimulations, volume of sound) were delivered directly from the automatic algorithm of the Dreem headband (Arnal et al., 2020) or were collected after manual scoring of the two PSGs. A correlational analysis was done between the absolute REM duration on average during one week at home and the stress variables (RMSSD, SUDS, ns-SCRs) at T3 for the two groups separately. The same analysis was done with the number of auditory stimulations on average during one week. Before the correlations were performed, a normality test was done on the different variables that were used, to use the appropriate test.

#### Emotional dream content

Among all emotions reported in the dreams, we focused on fear (see Introduction section), which was assessed as follows. We calculated the proportion of dreams containing fear in the week without the stimulations and during the week with stimulations. More specifically, we took the number of dreams where the emotion was reported as being present divided by the number of dreams where participants reported that they remembered the content of the dream. This gave us a proportion of this emotion for each participant, with one value for the week before stimulations and one value for the week with stimulations.

A correlational analysis was done between the change of fear in dreams (average of fear during the second week with stimulations minus the first week without stimulations) and the primary outcome measures (RMSSD, SUDS) and secondary outcome measure (ns-SCRs) at T3 for the two groups separately.

## 3. Results

Recruitment took place from July 2020 to June 2021. Figure S1 provides a CONSORT diagram of study participants (Supplementary Material).

### 3.1. Baseline characteristics

There were no differences between the two groups in age, social anxiety (assessed by the Liebowitz Social Anxiety Scale), sleep quality (assessed by the Pittsburgh Sleep Quality Index), anxiety (assessed by the Beck Anxiety Inventory II), depression (assessed by the Beck Depression Inventory II), severity of insomnia (assessed by the Insomnia Severity Index) and vigilance at T1 (assessed by PVT), as indicated in Table 1.

**Table 1.** Means, standard deviations, and comparison between the control and TMR group, of the age and the initial scores at the Liebowitz Social Anxiety Scale (LSAS), the Pittsburgh Sleep Quality Index (PSQI), the Beck Anxiety Inventory (BAI), the Beck Depression Inventory II (BDI II), the Insomnia Severity Index (ISI) and the psychomotor vigilance task (PVT) preceding the first VR session.

|  | TMR group<br>(N=24) | Control group<br>(N=24) | t(df) | p |
| --- | --- | --- | --- | --- |
| Age | 24.7 ± 5.78 | 24.12 ± 3.97 | 0.4 (40,74) | 0.68 |
| Liebowitz | 101.08 ± 12.1 | 100.29 ± 11.95 | 0.22 (45,99) | 0.82 |
| PSQI | 3.20 ± 1.28 | 3.08 ± 1.24 | 0.34 (45,96) | 0.73 |
| BDI | 7 ± 6.06 | 6.62 ± 5.41 | 0.22 (45,43) | 0.82 |
| BAI | 26.5 ± 14.85 | 21.16 ± 13.83 | 1.28 (45,76) | 0.2 |
| ISI | 3.83 ± 2.42 | 3.12 ± 2.32 | 1.03 (45,92) | 0.3 |
| PVT | 269.62 ± 36.03 | 263.83 ± 22.79 | 0.66 (38,87) | 0.5 |

Detailed results on the comparison between stress levels of the preparation, baseline and feedback phases of the VR sessions are reported in Supplementary Material (S3), while comparison of several sleep measures across time and groups are reported in Supplementary Material (S4).

### 3.2. Effects of TMR on social anxiety (T1a, T2, T3)

#### RMSSD

No significant group x time interaction was shown (F(2,76.848) = 0.7393, p = 0.481), nor a main effect of time (F(2,76.799) = 1.0190, p =0.366). Results showed a significant main effect of group (F(1,41.035) = 4.1536, p = 0.048). Follow-up pairwise comparisons indicated a significant difference between the groups during the preparation period T3 (t(92.2) = 2.119, p=0.037, Cohen’s d= 0.61), with lower RMSSD levels for the TMR group (M = 35.8, SD = 21.3) compared to the control group (M = 51.1, SD = 27.7). Moreover, a trend toward significant difference was observed between groups during the preparation period T2 (t(89.6) = 1.815, p = 0.075, Cohen’s d= 0.62), with lower RMSSD levels for the TMR group (M = 37.14, SD = 17.96) compared to the control group (M = 49.26, SD = 20.76).

#### SUDS

Analysis on SUDS scores showed no significant group x time interaction (F(2,88) = 0.0142, p = 0.986) and no main effect of group (F(1,44) = 0.3382, p = 0.564). There was a significant main effect of time, F(2,88) = 30.0611, p < 0.001). Follow-up pairwise comparison between time points revealed a significant difference between SUDS scores of T1a (M = 6.91, SD = 2.13) and T2 (M = 5.49, SD = 2.15) (t(88) = 5.53, p < 0.001), T1a and T3 (M = 4.98, SD = 2.49) (t(88) = 7.46, p < 0.001) and a trend towards a significant difference between SUDS scores of T2 and T3 (t(88) = 1.94, p = 0.056). The significant differences were observed for both groups.

#### ns-SCRs

No significant group x time interaction was shown (F(2, 71.174) = 0.3229, p = 0.725), nor main effect of group (F(1,40.772) = 0.1041, p = 0.749), nor main effect of time (F(2,71.120) = 0.0691, p = 0.933).

The results are illustrated in Figure 2 and the means, standard deviations per groups and p-values of the pairwise comparisons are reported in Table 2.

**Figure 2.**
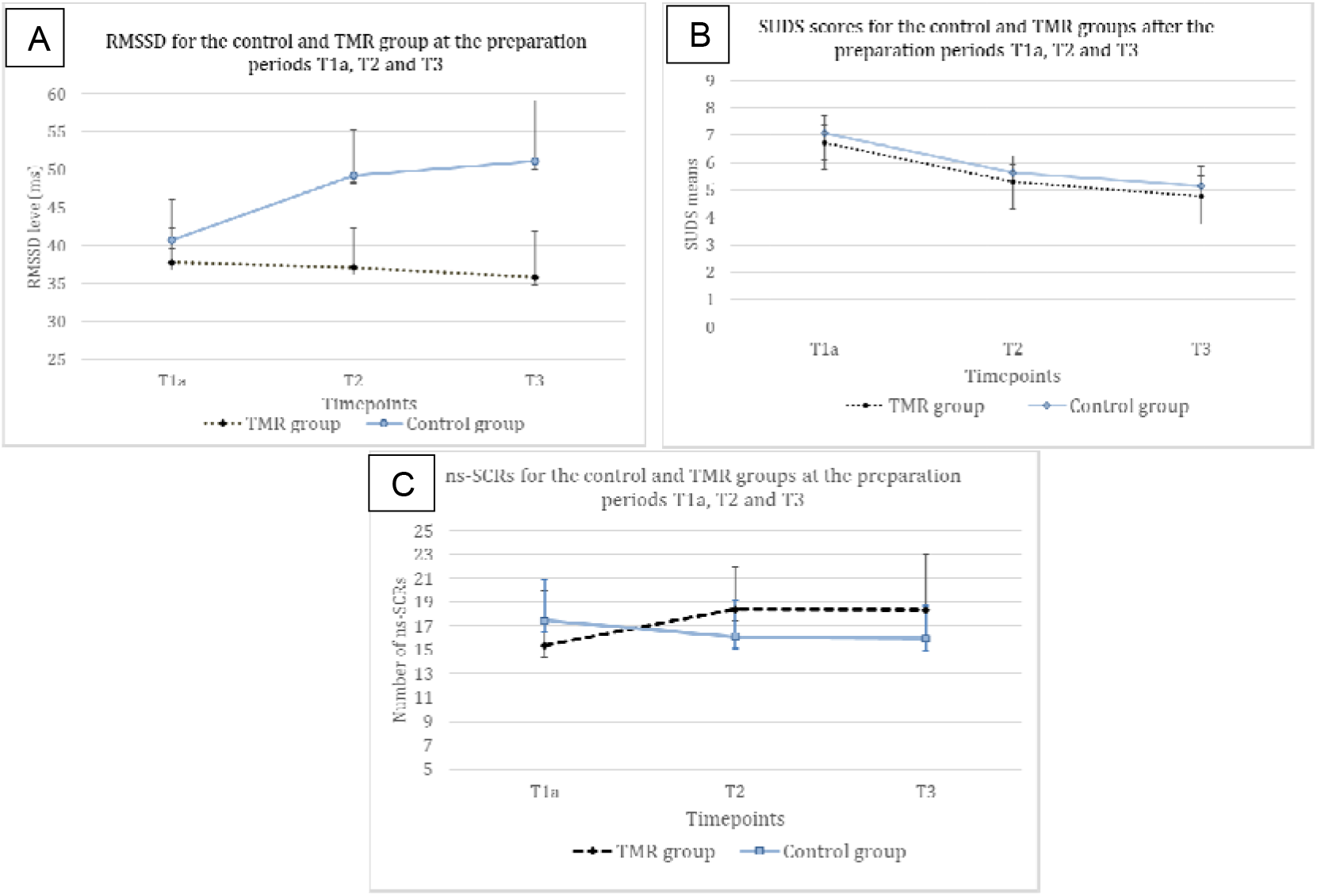
Mean values for the level of RMSSD (A), score of SUDS (B) and number of ns-SCRs (C) for the control and TMR groups during the preparation periods T1a, T2 and T3. Pairwise comparison showed a significant difference between the groups at T3 for RMSSD measure (p = 0.037). Regarding SUDS scores, there was a significant main effect of time (p < 0.01) and pairwise comparison between time points revealed a significant difference between SUDS scores of T1a and T2 (p < 0.001), T1a and T3 (p < 0.001) for each group. N = 46. Error bars represent 95% CI.

**Table 2.**
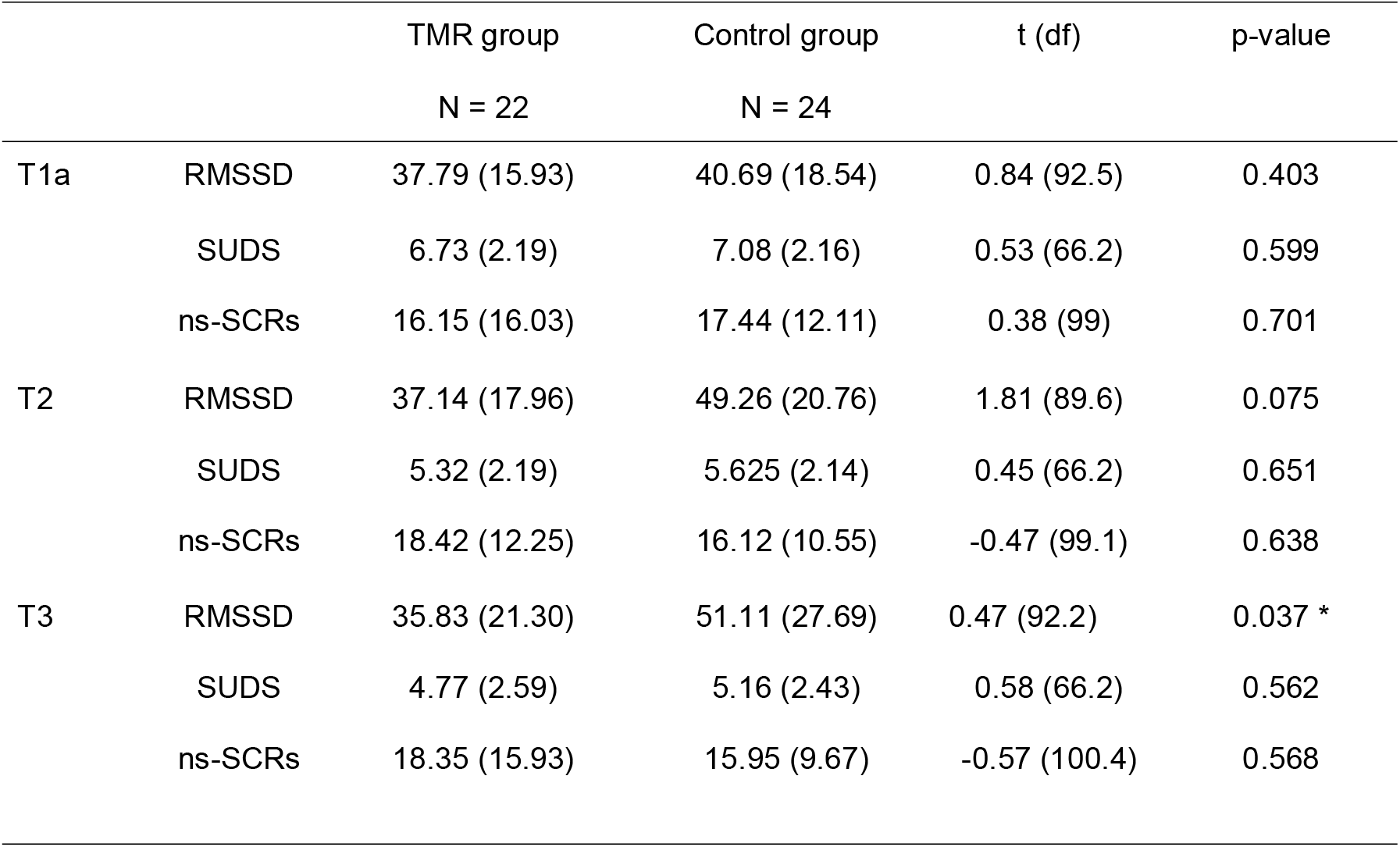
Means (standard deviations), and p-values (comparing groups) of the TMR and control groups on RMSSD (ms) levels, SUDS score, and ns-SCRs (number of events) during the preparation periods T1a, T2 and T3 preparation periods (* p < 0.05). N = 46.

|  |  | TMR group<br>N = 22 | Control group<br>N = 24 | t (df) | p-value |
| --- | --- | --- | --- | --- | --- |
| T1a | RMSSD | 37.79 (15.93) | 40.69 (18.54) | 0.84 (92.5) | 0.403 |
|  | SUDS | 6.73 (2.19) | 7.08 (2.16) | 0.53 (66.2) | 0.599 |
|  | ns-SCRs | 16.15 (16.03) | 17.44 (12.11) | 0.38 (99) | 0.701 |
| T2 | RMSSD | 37.14 (17.96) | 49.26 (20.76) | 1.81 (89.6) | 0.075 |
|  | SUDS | 5.32 (2.19) | 5.625 (2.14) | 0.45 (66.2) | 0.651 |
|  | ns-SCRs | 18.42 (12.25) | 16.12 (10.55) | -0.47 (99.1) | 0.638 |
| T3 | RMSSD | 35.83 (21.30) | 51.11 (27.69) | 0.47 (92.2) | 0.037 * |
|  | SUDS | 4.77 (2.59) | 5.16 (2.43) | 0.58 (66.2) | 0.562 |
|  | ns-SCRs | 18.35 (15.93) | 15.95 (9.67) | -0.57 (100.4) | 0.568 |

### 3.3. Effect of long-term TMR on generalization of anxiety (T0, T4)

#### RMSSD

Regarding RMSSD levels, results showed no group x time interaction (F(1, 37.405) = 0.0005, p = 0.982), no main effect of group (F(1, 39.291) = 0.5326, p = 0.469) and no main effect of time (F(1,37.405) = 0.7808, p = 0.382). No time effect was found for neither group and pairwise comparisons revealed no significant difference between groups at any time points

#### ns-SCRs

Analysis revealed no significant group x time interaction (F(1, 34.555) = 0.0846, p = 0.773), and no main effect of group (F(1, 35.903) = 0.2140, p = 0.646). A main effect of time was found (F(1, 34.416) = 6.5407, p = 0.015), indicating a higher number of ns-SCRs during the first generalization task (M = 12.9, SD = 8.5) compared to the last one (M = 8.7, SD = 6.1). This effect was trending toward significance for the control group (t(35.8) = 1.9637, p = 0.057) and not significant for the TMR group (t(33.2) 1.6617, p = 0.105). Pairwise comparisons revealed no significant differences between groups at any time points.

The results are illustrated in Figure 3 and the means, standard deviations per groups and p-values of the pairwise comparisons are reported in Table 3.

**Figure 3.**
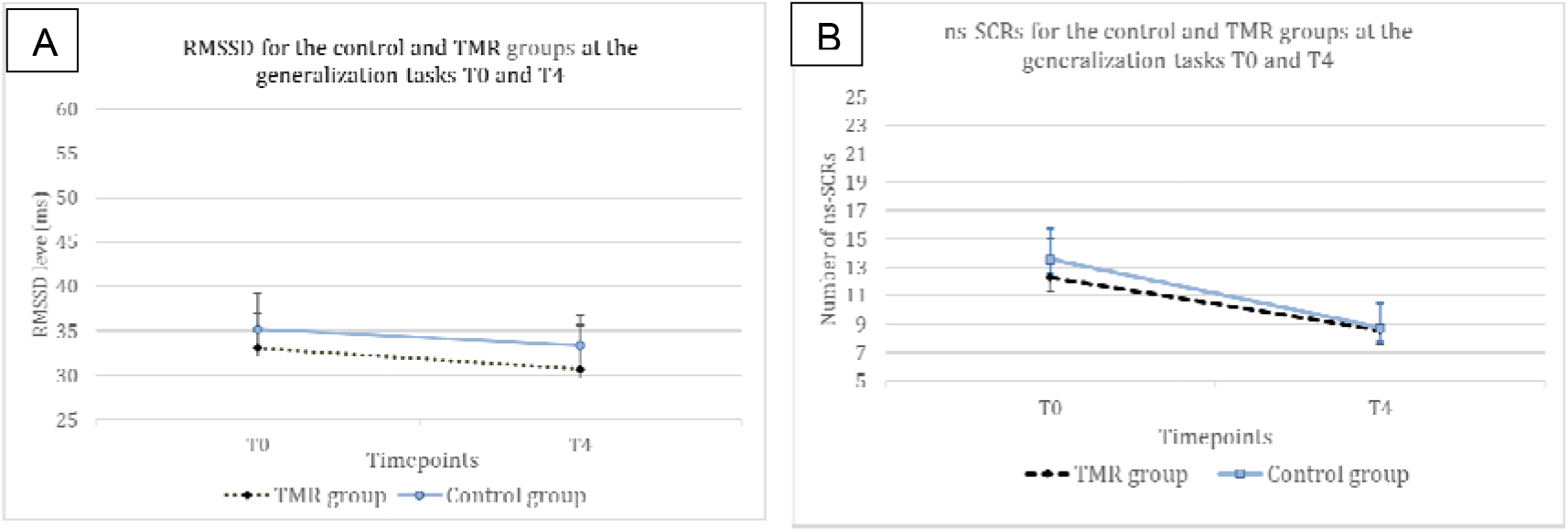
Means and standard deviation for the RMSSD levels (A) and the number of ns-SCRs (B) for the control and TMR groups during the generalization task periods T0 and T4. A type II ANOVA breakdown of fixed effects, following a multilevel model, showed a main effect of time on the number of ns-SCRs (p = 0.015) and with trend significance for the control group (p=0.057). N = 46. Error bars represent 95% CI.

**Table 3.** Means (standard deviation), and p-values (comparing groups) of the TMR and control groups on the ns-SCRs (number of events) and RMSSD (ms) levels during the generalization task periods (T0 and T4). N = 46.

|  |  | TMR group<br>N = 22 | Control group<br>N = 24 | t (df) | p-value |
| --- | --- | --- | --- | --- | --- |
| T0 | RMSSD | 33.09 (13.47) | 35.24 (14.13) | 0.56 (70.6) | 0.579 |
|  | ns-SCRs | 12.28 (9.59) | 13.57 (7.44) | 0.54(68.4) | 0.586 |
| T4 | RMSSD | 30.71 (17.11) | 33.42 (11.78) | 0.61 (68.8) | 0.5433 |
|  | ns-SCRs | 8.61 (6.64) | 8.75 (5.75) | 0.14 (69.3) | 0.889 |

### 3.4. Association between REM, TMR and stress levels

#### RMSSD

RMSSD levels in T3 correlated with REM duration in the TMR group (p=0.047, tau=0.307). The control group did not present this effect (p=0.916, tau=-0.019). We used a Kendall correlation as the distribution of RMSSD was not normal (p=0.001). After transformation of tau values to Pearson r-values (Walker, 2003), a Fisher r-to-z transformation test showed that the coefficients were significantly different (p=0.046, z=1.68).

A similar effect was observed for the correlation between the number of auditory stimulations and RMSSD in T3. The TMR group showed a significant effect (p=0.034, tau=0.164) but not the control group (p=0.5265, tau=-0.118). After transformation of tau values to Pearson r-values (Walker, 2003), a Fisher r-to-z transformation test showed that the coefficients were significantly different (p=0.003, z=2.75).

Importantly, this significant positive correlation between REM sleep duration/ auditory stimulations and RMSSD was not present after one night of stimulations (T2) and no significant correlation was found between the stress variables in T2 or T3 and the other sleep stages (N1, N2, N3).

#### SUDS

The correlation between REM duration and SUDS in T3 did not show any significant results for the TMR group (p=0.295, r=0.222) and the control group (p=0.368, r=-0.191).

No significant results were found for the correlation between the number of auditory stimulations and SUDS in T3, in the TMR group (p=0.522, r=0.137) and the control group (p=0.941, r=0.015).

#### ns-SCR

The correlation between REM duration and ns-SCR in T3 did not show any significant results for the TMR group (p=0.103, r=0.396) and the control group (p=0.546, r=-0.162).

Concerning the correlation of the number of auditory stimulations and ns-SCR in T3, no significant results were found in the TMR group (p=0.39, r=0.215) or the control group (p=0.441, r=-0.207).

There was also a positive correlation between the number of auditory stimulations and the duration of REM sleep over a week (r=0.5377, p=8.101e-05). A mediation test was done to understand if the number of auditory stimulations were a mediator to the effect of REM sleep on stress levels in T3. For this, we performed a bootstrap test and found that there was no significant result. Neither the average causal mediation effect (ACME) (p=0.77), nor the average direct effect (ADE) (p=0.38) or the total effect (p=0.39) were significant. These results indicate that the effect of auditory stimulations on stress levels is not mediated by the duration of REM sleep.

### 3.5. Link between fear in dreams and stress in wakefulness

No significant effect was found for the correlation between the change of fear in dreams (average of fear during the second week with stimulations minus the first week without stimulations) and RMSSD in T3 (p=0.1912, tau=-0.242) for the TMR group or the control group (p=0.791, tau=-0.044).

A significance is shown for the correlation between the change of fear and SUDS for the TMR group (p=0.0046, r=0.6513), but not for the control group (p=0.759, r=-0.0712). A Fisher r-to-z transformation test showed that the coefficients were significantly different (p=0.009, z=-2.382).

A similar significant correlation between the change of fear in dreams and ns-SRCs was found for the TMR group (p=0.0155, r=0.6308), but not for the control group (p=0.7995, r=-0.0747). A Fisher r-to-z transformation test showed that the coefficients were significantly different (p=0.011, z=-2.294).

## 4. Discussion

The primary hypothesis of this study was to test if adding TMR during REM sleep in addition to standard exposure therapy would be more effective than exposure therapy alone in reducing anxiety in individuals suffering from SAD. TMR consisted in associating a sound with an isolated extinction period (i.e., the positive feedback of exposure therapy), and reactivating this memory by presenting the same sound during REM sleep. Contrary to our hypothesis, we found that RMSSD was lower in the TMR group compared to the control group during the preparation period preceding a speech after 8 days of auditory stimulation during REM sleep in both groups (T3). No similar result was observed for the subjective measure (SUDS) nor the ns-SCRs at T3. We also found that subjective anxiety was reduced across the experiment for both groups (p < 0.001), indicating the efficiency of a small number of exposure therapy sessions in reducing social anxiety. In accordance with the depotentiation theory of REM sleep, secondary analyses also indicated that the number of auditory stimulations during REM sleep and REM sleep duration were negatively correlated with RMSSD in the TMR group. A positive correlation between increased presence of fear in dreams and SUDS/ns-SCRs was also observed in the TMR group, reflecting a possible link between emotional content in dreams and waking stress levels in these patients.

### 4.1. Effect of time

Among the three measures of anxiety, the SUDS was the only one to show an effect of time, indicating higher anxiety during the first preparation period (T1a) compared to the third (T2) and last (T3) ones in both groups. By contrast, the physiological measures did not show any modulation by time, probably because the small number of exposure therapy sessions in this protocol did not allow for such an improvement. The effect of time on subjective anxiety is in line with studies showing that exposure is efficient in reducing symptoms of anxiety disorders even without any other manipulation (Borgeat et al., 2009; Kaczkurkin & Foa, 2015; Klinger et al., 2005). However, strictly speaking, the lack of a control group without exposure therapy in the design of the current study cannot rule out that the significant reduction of subjective anxiety was not related to other factors (e.g., patient expectations; see limitations section below).

### 4.2. Challenges to isolate a safety memory

The comparison of the two groups on the primary psychophysiological outcome measure (RMSSD) showed a statistical significance at T3 for lower RMSSD levels in the TMR group compared to the control group during the preparation phase. No significant results between groups for the primary subjective measure of anxiety (SUDS) and the secondary psychophysiological outcome measure (ns-SCRs) were found at any time point. Therefore, TMR during REM sleep in our experiment did not enhance the beneficial effect of exposure therapy on anxiety-related distress (SUDS). Although RMSSD is not a specific measure for anxiety (i.e., it can be modified by other factors, see also Limitations), an explanation for these results could be that the sound in our experiment was not sufficiently associated with a safety memory (i.e., positive feedback). Indeed, when comparing anxiety levels of the preparation and feedback periods, physiological measures generally indicated similar levels between the two periods (Supplementary Material S3). Furthermore, anxiety levels were higher during feedbacks compared to the baseline periods, further demonstrating that the participants were still in a state of physiological stress during the feedback periods (Supplementary Material S3). Moreover, as patients with SAD show altered processing of feedback compared to healthy controls (Glazier & Alden, 2019; Koban et al., 2017; Voegler et al., 2019), positive feedback might not have been sufficient to change their expectations of a negative outcome.

Previous studies have associated a sound to aversive stimuli and presented this sound (conditioned stimulus, CS) during NREM sleep. This TMR procedure led to either a reduction in fear responses (Hauner et al., 2013; He et al., 2015) or increase of fear responses (Barnes & Wilson, 2014; Rolls et al., 2013). Protocol differences, such as reinforcement contingencies, relevance of the aversive stimuli or frequency of cueing during sleep, could explain such discrepancies (Diekelmann & Born, 2015). While these studies implicated memory reactivation during NREM sleep, and not REM sleep, these findings bring attention to the necessity to test multiple new experimental protocols in order to identify the mechanisms allowing sleep cueing to enhance extinction and those leading to opposite results. To this date, we do not know if consolidation of a positive memory during REM sleep would be more advantageous (Schwartz et al., 2022) than consolidation and subsequent depotentiation of a negative one (van der Helm et al., 2011).

### 4.3. Effect of REM sleep and TMR on stress

We here found a positive correlation between REM duration over one week and RMSSD in T3 in the TMR group. Although pertaining to a secondary analysis and limited by its correlational nature, this result in a clinical population (i.e., SAD patients) supports the idea that REM sleep is beneficial for reducing some physiological manifestations of stress levels during wakefulness, in line with enhanced extinction (Menz et al., 2016; Pace-Schott et al., 2014; Spoormaker et al., 2012) and emotional depotentiation (van der Helm et al., 2011; Walker & van der Helm, 2009). Importantly, this correlation was not found for other sleep stages, in accordance with previous research (Gujar et al., 2011; Menz et al., 2016; Rihm & Rasch, 2015; van der Helm et al., 2011; Walker & van der Helm, 2009). Indeed, results on the possible effects of N2 sleep and/or SWS on extinction learning are inconsistent. Some results show that N2 sleep is beneficial to reduce self-reported fear for spider phobia (Kleim et al., 2014), while others show that N2/SWS (with or without TMR) does not enhance exposure therapy for spider phobia (Rihm et al., 2016) or for SAD (Pace-Schott et al., 2018). As previously mentioned, TMR during SWS can also either enhance extinction (Hauner et al., 2013; He et al., 2015) or strengthen a fear memory (Ai & Dai, 2018; Barnes & Wilson, 2014; Rolls et al., 2013). Thus, REM sleep would not strengthen the emotional tone of memories (Baran et al., 2012; Jones & Spencer, 2019; Wagner et al., 2002; Werner et al., 2015), but might instead consolidate negative memories while attenuating their emotional tone (Cunningham et al., 2014; Gujar et al., 2011; Rihm & Rasch, 2015; van der Helm et al., 2011; Walker & van der Helm, 2009). REM sleep and dreaming (Sterpenich et al., 2020; Vallat et al., 2017) might offer a permissive condition for the remodeling of negative experienced events.

Previous studies have explored the emotional role of REM sleep mainly in healthy participants. Our study suggests that REM sleep could benefit anxious patients but only after a TMR manipulation. Indeed, a positive effect of REM sleep on stress reduction was only seen in the TMR group of our study, where the sound has been associated with a previous waking emotional event. This observation is consistent with previous studies showing that REM sleep enhanced extinction learning only after an extinction training took place before sleep (Fu et al., 2007; Menz et al., 2016; Silvestri, 2005). Notably, the aforementioned beneficial effects of TMR during REM sleep did not appear after one single night, but only after one week of stimulation at home. Repeated stimulation over successive nights might be needed to permanently consolidate the formation of a new (i.e. initially labile) safety memory during sleep (e.g. make it hippocampus-independent (Frankland & Bontempi, 2005)).

### 4.4. Association between fear in dreams and anxiety in SAD patients

The frequency of fear experienced in dreams correlated with the primary clinical outcome measure (SUDS) and the secondary psychophysiological outcome measure (nd-SCRs) in the TMR group (but not in the control group). The more these participants experienced fear in their dreams, the more they were stressed in T3, as measured with nd-SCRs and SUDS. In a previous study (Sterpenich et al., 2020), we demonstrated that increased fear in dreams benefits reduced stress when healthy participants are exposed to fearful stimuli during wakefulness, and which was in accordance with an extinction function of dreaming in healthy participants (Nielsen & Levin, 2007). The results of the present study suggest that this fear extinction function of dreams (Nielsen & Levin, 2007; Sterpenich et al., 2020) might be deficient in clinical populations, such as anxious people here. For example, although healthy participants experiencing fearful dreams have higher mPFC activity in fearful situations in wakefulness (Sterpenich et al., 2020), nightmare patients demonstrate a decreased mPFC activity during the viewing of negative pictures (Marquis et al., 2016). We speculate that a similar failure of the fear extinction function of dreams can explain our result in patients with SAD, who also demonstrate a decreased mPFC activity and increased amygdala activity during social stressors (Marazziti et al., 2015). Whether there is a causal relationship between anxiety disorders and such a deficient extinction function of dreaming in these patients should be tested in future studies.

### 4.5 Limitations

Apart from the difficulty to isolate a safety memory (see section 4.1), there are some other limitations in this study. First, all three measures of anxiety did not always concur. On one hand, a dissociation between physiological and subjective measures of anxiety is very common (Campbell & Ehlert, 2012; Pace-Schott et al., 2018) and seems to reflect a different response of the autonomic nervous system and of subjective (conscious) experience to perceived stress (Grillon et al., 2009 ; Pace-Schott et al. 2018). On the other hand, this dissociation may be related to the use of a VR setting for the measurement of stress during exposure therapy. While such a setting allows for realistic and controlled exposure to feared stimuli, it may have reduced the participants’ immersion in the feared situation (Kritikos et al., 2019; Wilhelm et al., 2005b). Comparing subjective assessments with different physiological measurements is needed to have more insight on the implication of different systems in stress responses (Kerous et al., 2020). Another limitation, linked to the assessment of anxiety, relates to RMSSD, as one of our principal results concerns RMSSD levels, which were lower for the TMR group at T3. While in our study we use RMSSD to assess anxiety levels (Chalmers et al., 2014), the fact that it can be modulated by other factors should not be overlooked. Indeed, RMSSD reflects some physiological manifestations of stress, but has also been linked to depression, worry, measures of inflammation or cognitive functions involving the prefrontal cortex (McCraty & Shaffer, 2015).

Moreover, it is possible that a reduction of sleep quality/quantity may have accounted for the negative results of our study. Indeed, total sleep time (TST) and N2 sleep were significantly decreased in the TMR group during the week at home compared to the habituation night (see Supplementary Material S4, Table S1). Moreover, TMR participants were almost significantly (p=.08) less vigilant compared to those of the control group, as measured by the PVT before the last VR session at T3 (see Supplementary Material S4, Table S1), while HRV is sensitive to sleep deprivation (Bourdillon et al., 2021). This means that, even though REM sleep and auditory stimulations had a positive effect on stress within this group, sleep quality/quantity may be a more important determinant for the participant’s well-being and stress levels (although the relationship between stress and sleep quality/quantity is bidirectional) (Lo Martire et al., 2020). Indeed, TMR seems to lose its positive effect when sleep quality/quantity is compromised (Göldi & Rasch, 2019), while sleep quality, which is already poor in SAD patients compared to healthy controls (Horenstein et al., 2019), predicts treatment outcome of exposure therapy in patients with SAD (Kushnir et al., 2014; Zalta et al., 2013). Unfortunately, we could not establish whether the decrease of TST in the TMR group resulted from TMR itself, as the between-group comparisons for TST during the week at home and/or for the last night before T3, did not yield any significant results. Yet, we suspect that the decrease of N2 sleep is related to the application of auditory stimulations, as it is also observed in the control group (which however does not present a significant decrease in TST). Our results hence suggest that the strengthening of an extinction memory during REM sleep with TMR can be helpful to reduce stress only when controlling for sleep disruption, in line with previous research (Göldi & Rasch, 2019). Indeed, in a recent paper (Davidson & Pace-Schott, 2020), the authors stated that the beneficial effects of REM sleep on extinction are not noticeable if sleep is disturbed.

Finally, this experiment lacks a SAD group without any stimulation or a SAD group without exposure therapy. Therefore, although we know that exposure therapy promotes extinction (Craske et al., 2018) and that REM sleep enhances extinction in healthy participants after an extinction task compared to wakefulness or NREM sleep (Menz et al., 2016; Pace-Schott et al., 2014), we cannot safely conclude from our study that TMR during REM sleep (even without sleep disruption) would add an extra benefit in extinction processes. Indeed, previous studies indicate the presence of a ceiling effect of the highly effective exposure therapy or of REM sleep alone in extinction processes (Pace-Schott et al., 2014; Rihm et al., 2016).

As a technical note, in the present study, we applied TMR during REM sleep, irrespective of REM tonic or phasic states. Yet, for the purpose of extinction consolidation, TMR should ideally be performed during phasic REM sleep, as phasic P-waves seem important for the retention of fear extinction memory occurring after the acquisition of fear extinction learning (Datta & O’Malley, 2013). Therefore, future technical development of TMR applied during REM may aim at achieving selective stimulations during phasic REM.

### 4.6 Future perspectives

To the best of our knowledge, this is the first TMR study exploring the links between REM sleep, extinction and anxiety in a clinical population. Although this study does not allow to ascertain a positive effect of TMR on SAD, TMR remains an efficient tool for the reprocessing of emotional memories (Rihm & Rasch, 2015; Sterpenich et al., 2014), and further research with TMR could be the key to develop efficient therapies for SAD or other emotional disorders implicating deficient extinction learning. TMR application during sleep may avoid certain disadvantages of traditional exposure therapies during wakefulness, such as worsening mood and anxiety during the recall of painful experiences. By deploying and popularizing easy-to-use devices at home in order to enhance the consolidation of safety memories, these therapies could reach a big part of the general population and lead to new approaches for promoting emotional well-being.

Future studies should better characterize the nature of the link between anxiety disorders and the deficient extinction function of dreaming. They could also test whether Imagery Rehearsal Therapy (IRT), an efficient treatment for reducing negative emotions in dreams, and specifically nightmares (Krakow & Zadra, 2010), can be helpful for treating anxiety disorders as well. In this direction, it has been already shown that IRT in PTSD patients can improve not only nightmare frequency and intensity, but daytime symptoms too (PTSD symptoms, depression) (Krakow et al., 2001).

## Data Availability

All data produced in the present study are available upon reasonable request to the authors.

## Author Contributions

SS and LP designed the experiments, FB, PH, FG, CP and LP conducted the experiments, FB, PH, FG, CP and LP analyzed the data, FB, PH, FG, CP, SD, SS and LP wrote the paper.

## Acknowledgments

The authors would like to thank Virginie Sterpenich, Guillaume Legendre, Kinga Igloi, Guido Bondolfi, Paolo Cordera, Melanie Schaffer, Gwenael Birot, Margaux Dubessy, Stephen Perrig, Ralph Schmidt for their useful help and suggestions for this study. This study was supported by the Human Neuroscience Platform, Fondation Campus Biotech Geneva, Geneva, Switzerland. Clinical trial registration: Clinicaltrials.gov Identifier NCT05261659.

