## Supplementary Material for "Targeted memory reactivation during REM sleep in patients with social anxiety disorder"

**S1. Analysis of psychophysiological measures**

***Analysis of electrocardiogram signal (ECG)***
Electrocardiogram (ECG) signal was recorded during the VR sessions and during the nights spent at the Campus Biotech. The signal was measured using disposable adhesive surface snap hydrogel electrodes, with a diameter of 24mm (reference: CDES000024, Spes Medica S.r.l) or 20x25mm (DENIS01520, Spes Medical S.r.l.). ECG measures were recorded wirelessly using the Biopac MP160 System, (Biopac Systems Inc., Goleta, CA, 2013) and the software AcqKnowledge v.5.0. ECG signal was recorded using the V-Amp (Brain Products, Gilching, Germany), and analyzed with the Kubios HRV analysis software. Eventual artefacts were detected and corrected manually and/or by using the automatic medium correction provided by the software. When needed, the following filters were applied: Low-Pass filter (Blackman -92dB) fixed at 30Hz (133 coefficients); High-Pass filter (Blackman -92dB fixed at 8Hz (800 coefficients). For our study, we were interested in the root mean square of successive differences between normal heartbeats (RMSSD, in ms).

***Analysis of electrodermal activity (EDA)***
EDA was recorded during the VR sessions using two disposable Ag/AgCl snap electrodes (EL507, manufactured for BIOPAC Systems, Inc.), pre-gelled with isotonic gel (size: 27mm wide x 36mm long x 1.5 mm thick). The signal was analyzed with the software AcqKnowledge v.4.4. The data signal was first filtered with a Low-Pass filter (Blackman -61 dB, cutoff frequency = 1 Hz), then downsampled to 100Hz. Phasic and tonic EDA were derived from the signal using High-Pass (Blackman -61 dB, cutoff frequency = 0.05 Hz) and Low-Pass (Blackman -61 dB, cutoff frequency = 0.05 Hz) filters respectively, in order to obtain skin conductance responses (SCRs) and skin conductance levels (SCLs). For our study, we were interested in non-specific SCR (ns-SCRs), which were located with a threshold level 0.02μS. The number of events of ns-SCRs were counted for each VR session phase. Eventual artefacts were visually detected and corrected. The higher is the number of ns-SCRs, the more the individual can be considered stressed (Braithwaite et al., 2013) .

**S2. Detailed description of the experimental procedure**

### ***Before the experimental sessions*** After completing online questionnaires (on demographic and medical information, sleep quality, anxiety and depression levels), participants were given an actigraph to wear, and a sleep diary and sleep agenda to fill, one week before the habituation night. The duration of the experiment was ten days and was organized in the following way: habituation night at the lab (Day 0), a VR session and an experimental night at the lab (Day1), a VR session in the morning (Day 2), one week sleeping at home (Day 2-8), a VR session in the morning of Day 9.

***Day 0 - Habituation night***
Participants spent a night at the lab in order to get used to the polysomnography and the Dreem headband setup. Subjects arrived at the laboratory at 21:00 and the electrodes for the polysomnography and ECG recordings, as well as the Dreem® headband were placed. The participants then went to bed between 23:00 and 24:00. There was no stimulation (with the sound) during the habituation night. The next morning, after waking up, the participants filled a post-sleep questionnaire.

### ***Day 1 – VR session and Experimental night*** Participants arrived at 20:00 and did a PVT task. They participated in the first VR session. The aim was to expose participants to different social contexts and create an exposure therapy while measuring their stress level. This VR session was divided into the following phases. - *Baseline*. A three-minute phase during which participants had to stay seated, relax and get used to the environment. The goal of this rest phase was to allow any distress the participant might feel to dissipate. - *Generalization task (T0)*. A three-minute and 50 seconds phase. Participants were instructed to stand on a red square, not to move their body, but could move their heads to look around them. Virtual characters could be seen discussing among themselves, before progressively gathering around the participant to look at them. The goal of the generalization task is to investigate the generalization of extinction. - *Baseline* - *Preparation (T1a)*. Participants were given a topic and asked to prepare a presentation for five minutes. They were instructed to stay seated and were not allowed to use any material. A timer allowed them to monitor the remaining preparation time. The topic was: *“Tell us about topics in the news that struck you and why”*. At the end of the preparation, the curtains leading to the stage opened. - *Presentation (T1a)*. A 5-minutes period during which participants made their presentation, while standing in front of the virtual microphone. - *Feedback (T1a)*. This phase lasted 3 minutes and 30 seconds (approximately). Participants received positive feedback composed of three parts. First, a 10 second applause by the virtual public and jury. Then 10 to 20 seconds of feedback from both members of the jury, which was positive in the content of their speeches (e.g., *“Thank you very much for your speech. We can see that you made a good effort. We know that the task was not easy, but you were very devoted to it”*) and through positive facial expressions and gestures. Finally, the experimenter gave a feedback of about 3 minutes to the participants. Each feedback made by the experimenter was personalized according to each presentation of each subject, but mainly consisted of comments on the clarity, organization, content, duration, and relevance of the presentation. During this phase, a 1-second sound (i.e., a piano chord) was played every 10 seconds through a headphone for the participants in the TMR group, in order for an association between this positive feedback experience (representing an extinction memory) and the sound to take place. After the feedback, the curtains closed, and the participants were instructed to sit back down on the chair backstage. - *Baseline* - *Preparation (T1b)*. The topic was: *“Present your job and/or studies and why you have chosen that”* - *Presentation (T1b)* - *Feedback (T1b)*

Following this VR session, the participants spent the night at the lab with the polysomnography and Dreem headband setting. Explanations regarding how to use the headband and app were made. During the experimental night, the sound was presented during REM sleep for both groups.

### ***Day 2 – VR session***

### The next morning participants filled a post-sleep questionnaire. Participants did a five minute PVT and then took part in the VR session T2, composed of the following periods: - *Baseline* - *Preparation (T2)*. The topic was: *“Present yourself as you would do for a job interview”.* - *Presentation (T2)* - *Feedback (T2)*

After that, if needed, they were shown how to use the Dreem® headband and mobile application again. Finally, they were given the headband, a phone with the Dreem® application, the instructions on how to use them, and they were reminded to continue to wear the actigraph and to fill in the dream diary and sleep agenda during the week at home.

***Day 3 to 8 - week at home***
During the week at home, the participants were instructed to wear the Dreem® headband every night, to fill the sleep diary and sleep agenda and to wear the actigraph. The experimenter was able to check that the stimulations were activated during the nights.

### ***Day 9 – VR session*** On the last day of the experiment, participants arrived at the lab between 10:00 and 11:00. They were administered a five-minute psychomotor vigilance task (PVT), the post-week at home questionnaire and they took part in the last VR sessions composed as follow: - *Baseline*. - *Preparation (T3)*. The topic was: *“Present your favourite book, movie or TV show and explain why you like it so much”.* - *Presentation (T3)* - *Feedback (T3)* - *Baseline* - *Generalization task (T4).*

**S3. Comparison of anxiety measures during baseline, preparation and feedback across the sessions of the main task (T1a, T2, T3) and the generalization tasks (T0, T4)**

Results of the multilevel analysis and follow-up t-tests with RMSSD and ns-SCR as dependent variables are reported. The RMSSD levels per minute and the number of ns-SCRs per minute (and not the total number of events during the entire period) were used (as the baseline, preparation and feedback periods were not of the same duration). We compared the RMSSD levels and the number of ns-SCRs levels between the main tasks (T1a, T2 and T3) and their respective baseline and feedback periods. We compared RMSSD levels and the number of ns-SCRs between the generalization tasks (T0 and T4) and their respective baseline periods.

***Baseline T1 vs Preparation T1a***
**RMSSD.** Regarding RMSSD levels, results showed a significant difference between baseline (*M* = 16.17, *SD* = 7.19) and preparation (*M* = 7.87, *SD* = 3.98), with lower levels during the preparation period, *t*(68.1) = 8.8411, *p* < 0.001. This difference was significant for both the TMR and the control group (*ps* < 0.01).
**ns-SCRs.** Results showed a significant difference between the first baseline (*M =* 1.31, *SD* = 1,27) and the first preparation period (*M* = 3.34, *SD* = 2.82), indicating that participants’ number of ns-SCRs were lower during the baseline period, *t*(70.4) = -7.1229, *p* < 0.01). This difference was significant for both the TMR and the control group (*ps* < 0.01).

***Baseline T1 vs Feedback T1*	RMSSD.** Results showed that the RMSSD levels were lower during feedback (*M* = 7.94, *SD* = 3.98) than baseline (*M* = 16.17, *SD* = 7.19), *t*(69) = 8.3616, *p* < 0.001. This difference was significant for both the TMR and the control group (*ps* < 0.01).
**ns-SCRs.** Results showed that the number of ns-SCRs was higher during feedback (*M* = 2.35, *SD* = 1.93) than baseline (*M =* 1.31, *SD* = 1,27), *t*(72.6) = -3.6357, *p* < 0.001. This difference was significant for the control group (*p* = 0.002) and trending towards significance the TMR group (*p* = 0.056).

***Preparation T1a vs Feedback T1*	RMSSD.** Results showed no significant difference in RMSSD levels between the feedback (*M* = 7.94, *SD* = 3.98) and preparation (*M* = 7.87, *SD* = 3.98) periods, *t*(68) = 0.1448, *p* = 0.88. This difference was not significant for either group.
**ns-SCRs.** Results showed that the number of ns-SCRs was lower during feedback (*M* = 2.35, *SD* = 1.93) than preparation (*M* = 3.34, *SD* = 2.82), *t*(73.3) = -3.3168, *p* < 0.001. This difference was significant for the TMR group (*p* = 0.011) and the control group (*p* = 0.042).

***Baseline T2 vs Preparation T2*
RMSSD.** Regarding RMSSD levels, results showed a significant difference between baseline (*M* = 16.97, *SD* = 8.24) and preparation (*M* = 8.67, *SD* = 4.03), with lower levels during the preparation period, *t*(74.9) = 10.5896, *p* < 0.001. This difference was significant for both the TMR and the control group (*ps* < 0.001).
**ns-SCRs.** Results showed a significant difference between the baseline T2 (*M* = 2.05, *SD* = 1.56) and the preparation period T2 (*M* = 3.51, *SD* = 2.24), indicating that participants’ number of ns-SCRs were lower during the baseline period, *t*(64.6) = -5.4543, *p* < 0.01). This difference was significant for both the TMR (*p* < 0.01) and the control group (*p* = 0.012).

***Baseline T2 vs Feedback T2***
**RMSSD.** Results showed that the RMSSD levels were lower during feedback (*M* = 7.49, *SD* = 3.22) than baseline (*M* = 16.97, *SD* = 8.24), *t*(75) = 12.2009, *p* < 0.001. This difference was significant for both the TMR and the control group (*ps* < 0.01).
**ns-SCRs.** Results showed that the number of ns-SCRs was higher during feedback (*M* = 3.48, *SD* = 2.43) than baseline (*M* = 2.05, *SD* = 1.56) *t(*66.7) = -5.3104, *p* < 0.001. This difference was significant for the control group (*p* = 0.003) and the TMR group (*p* < 0.001).

***Preparation T2 vs Feedback T2***

**RMSSD.** Results showed no significant difference in RMSSD levels between the feedback (*M* = 7.49, *SD* = 3.22) and preparation (*M* = 8.67, *SD* = 4.03) periods, *t*(74.6) = -1.8474, *p* = 0.068. This difference was not significant for either group.
**ns-SCRs.** Results showed no significant difference in the number of ns-SCRs between the feedback (*M* = 3.48, *SD* = 2.43) and preparation (*M* = 3.51, *SD* = 2.24) periods, *t*(67.5) = -0.1233, *p* = 0.902). This difference was not significant for either group.

***Baseline T3 vs Preparation T3*
RMSSD**. Regarding RMSSD levels, results showed a significant difference between baseline (*M* = 14.92, *SD* = 7.17) and preparation (*M* = 8.77, *SD* = 5.14), with lower levels during the preparation period, *t*(71.1) = 9.3316, *p* < 0.001. This difference was significant for both the TMR (*p* = 0.028) and the control group (*p* < 0.001). **ns-SCRs***.* Results showed a significant difference between the baseline T3 (*M =* 1.35, *SD* = 1.43) and the preparation period T3 (*M* = 3.5, *SD* = 2.17), indicating that participants’ number of ns-SCRs were lower during the baseline period, *t*(61.4) = -5.4871 , *p* < 0.01). This difference was significant for both the TMR (*p* = 0.005) and the control group (*p* = 0.005).

***Baseline T3 vs Feedback T3***
**RMSSD***.* Results showed that the RMSSD levels were lower during feedback (*M* = 7.53, *SD* = 3.11) than baseline (*M* = 14.92, *SD* = 7.17), *t*(70.9) = 9.4046, *p* < 0.001. This difference was significant for both the TMR and the control group (*ps* < 0.01).
**ns-SCRs.** Results showed that the number of ns-SCRs was higher during feedback (*M* = 2.86, *SD* = 2.08) than baseline (*M =* 1.35, *SD* = 1.43), *t*(61.3) = -4.4776, p < 0.001. This difference was significant for the control group (*p* = 0.004) and the TMR group (*ps* < 0.001).

***Preparation T3 vs Feedback T3***
**RMSSD.** Results showed no significant difference in RMSSD levels between the feedback (*M* = 7.53, *SD* = 3.11) and preparation (*M* = 8.77, *SD* = 5.14) periods, *t*(71.5) = -0.4618, *p* = 0.645. This difference was not significant for either group.
**ns-SCRs.** Results showed no significant difference in the number of ns-SCRs between the feedback (*M* = 2.86, *SD* = 2.08) and preparation (*M* = 3.5, *SD* = 2.17) periods, *t*(63) = -1.1130 , *p* = 0.269). This difference was not significant for either group.

***Baseline T0 vs Generalization T0*	RMSSD.** Results showed a significant difference between the baseline preceding T0 (*M =* 14.279, *SD* = 5.559) and the generalization task T0 (*M* = 9.104, *SD* = 3.638), indicating that participants’ RMSSD levels were higher during the baseline period, *t*(26.8) = 6.5130, *p* < 0.001). This difference was significant for both the TMR and the control group (ps < 0.001). **ns-SCRs.** Results showed a significant difference between the baseline preceding T0 (*M =* 2.69, *SD* = 1.92) and the first generalization task (*M* = 3.44, *SD* = 2.28), indicating that participants’ number of ns-SCRs were lower during the baseline period, *t*(36.7) = -2.5960, *p* = 0.01349). This difference was trending toward significance for both the TMR (p = 0.09311) and the control group (p = 0.05987).

***Baseline T4 vs Generalization T4*	RMSSD.** Results showed a significant difference between the baseline preceding T4 (*M =* 15.366, *SD* = 7.278) and the generalization task T4 (*M* = 8.561, *SD* = 3.866), indicating that participants’ RMSSD levels were higher during the baseline period, *t*(29.4) = 7.7417, *p* < 0.001). This difference was significant for both the TMR and the control group (ps < 0.001).	**ns-SCRs.** Results showed a significant difference between the baseline preceding T4 (*M =* 1.574, *SD* = 1.035) and the last generalization task (*M* = 2.314, *SD* = 1.639), indicating that participants’ number of ns-SCRs were lower during the baseline period, *t*(25) = -3.0731, *p* = 0.005). This difference was significant for the TMR group (p = 0.018) and trending toward significance for the control group (p = 0.078).

**S4.Sleep structure of participants**

Table S1 shows the values of several sleep parameters across time and groups. For several sleep variables (duration N2, N3, REM sleep, TST, REM sleep fragmentation), we performed mixed ANOVAs with “*Group*” (control vs TMR) as a between-subject factor and “*Time*” (habituation night vs experimental night vs week nights) as a within-subject factor.

***N2 stage***

We did not find any group (p=0.657, F=0.198) or a group*time interaction (p=0.777, F=0.08) effects.

We found a significant time effect (p=3.81e-07, F=28.466). Follow-up t-tests show a decrease of N2 sleep between the habituation night and the experimental night (p=0.0001) and between the habituation night and the week nights (p=1.068e-05) but not between the experimental night and the rest of the week (p=0.4828) for the TMR group. For the control group, the difference between the habituation night and the experimental night is not significant (p=0.05458) and becomes significant when compared to the rest of the week (p=0.00154).

***N3 stage***

We did not find any group (p=0.324, F=0.911), time (p=0.305, F=1.059) or group*time interaction (p=0.684, F=0.167) effects.

***REM stage***

We did not find any time (p=0.4806, F=0.5) or group*time interaction (p=0.7644, F=0.09) effects. There was a group effect (p=0.0368, F=4.447). A follow-up t-test showed a tendency for significance (p=0.08) for the TMR group to have more REM sleep during the week compared to the control group.

### ***Total sleep time***

We did not find any group (p=0.4842, F=0.492) or group*time interaction (p=0.7795, F=0.079) effects.

There is a significant time effect (p=0.0148, F=6.098). Follow-up t-tests show a decrease of TST between the habituation night and the experimental night (p=0.0008) and the rest of the week (p=0.008) in the TMR group, but no difference between the experimental night and the rest of the week (p=0.9127). For the control group, there are no differences between the habituation night and the experimental night (p=0.3594) or between the habituation night and the rest of the week (p=0.1478). There is also no significant difference between the experimental night and the rest of the week (p=0.3486).

***Fragmentation of REM sleep***

In order to test for the effects of auditory stimulations (associated or not) in sleep, we calculated a REM fragmentation score (REM_fragM =_ the number of awakenings during REM sleep for each night of each participant x 60 / REM duration of that night).

This score does not show any group (p=0.304, F=1.066), time (p=0.180, F=1.815) or group*time interaction (p=0.638, F=0.222) effects.

***Fear emotion in dreams***

The presence of the emotion of fear in dreams between the first week (without auditory stimulation and the second week (with auditory stimulations) does not show any group (p=0.743, F=0.108), time (p=0.181, F=1.824) or group*time interaction (p=0.813, F=0.056) effects.

Table S1. *Means, standard deviations, and comparison between groups of different sleep stages (N2, N3, REM) in minutes, total sleep time (TST) in minutes, volume of sound, number of stimulations, REM sleep fragmentation (REM_fragM =_ the number of awakenings during REM sleep x 60 / REM duration of that night) for the habituation night, experimental night and 8 nights with auditory stimulations at home, and average proportion average of fear in dreams before and during auditory stimulations.*

|  | TMR group | Control group | df | t | pvalue |
| --- | --- | --- | --- | --- | --- |
| REM habituation night | 133.63 ± 46.27 | 112.73 ± 52.95 | 43.24 | -1.42 | 0.16 |
| REM experimental night | 124.06 ± 46.06 | 114.64 ± 47.84 | 45.93 | -0.69 | 0.49 |
| REM week nights | 137.06 ± 28.59 | 121.61 ± 31.25 | 45.64 | -1.78 | 0.08 |
| N2 habituation night | 242.34 ± 54.69 | 235.65 ± 56.83 | 43.93 | -0.4 | 0.68 |
| N2 experimental nights | 192.7 ± 47.24 | 210.47 ± 49.92 | 45.886 | 1.26 | 0.21 |
| N2 week nights | 186.8 ± 38.66 | 185.99 ± 33.42 | 45.05 | -0.07 | 0.93 |
| N3 habituation night | 86.04 ± 36.63 | 97.43 ± 32.05 | 43.23 | 1.12 | 0.26 |
| N3 experimental night | 97.8 ± 44.65 | 97.47 ± 32.31 | 41.9 | -0.02 | 0.97 |
| N3 week nights | 96.22 ± 31.97 | 101.76 ± 24.13 | 42.79 | 0.67 | 0.5 |
| TST habituation night | 466.26 ± 80.22 | 445.06 ± 101.82 | 41.71 | -0.78 | 0.43 |
| TST experimental night | 420.45 ± 75.44 | 425.65 ±81.08 | 45.76 | 0.22 | 0.82 |
| TST week nights | 421.87 ± 65.96 | 409.97 ± 55.58 | 44.71 | -0.67 | 0.5 |
| Volume of sound week nights | 38.59 ± 22.39 | 40.76 ± 21.32 | 45.89 | 0.34 | 0.73 |
| Number of stimulations experimental night | 169.04 ± 106.48 | 175.54 ± 132 | 44.06 | -0.18 | 0.85 |
| Number of stimulations week nights | 164.48 ± 75.95 | 158.91 ± 84.62 | 45.47 | 0.23 | 0.81 |
| REM_fragm_ habituation night | 11.11 ± 28.39 | 7.28 ± 9.57 | 27.58 | -0.6 | 0.54 |
| REM_fragm_ experimental night | 6.4 ± 6.45 | 4.76 ± 4.4 | 40.6 | 1.02 | 0.31 |
| REM_fragm_ week nights | 6.31 ± 5.67 | 4.96 ± 3.97 | 41.2 | 0.95 | 0.34 |
| PVT 2 | 258 ±26.5 | 249 ± 29.8 | 46 | 1.03 | 0.3 |
| PVT 3 | 268.86 ± 45.2 | 248.62 ± 30.24 | 38.19 | -1.79 | 0.08 |
| Fear during week without stimulations | 0.33 ± 0.24 | 0.32 ± 0.34 | 43.64 | -0.08 | 0.92 |
| Fear during week with stimulations | 0.25 ± 0.32 | 0.21 ± 0.37 | 36.8 | -0.36 | 0.71 |

**Figure S1. Consort Diagram of the study**

Analysed (n= 22 )
♦ Excluded from analysis (unusable data) (n= 2)

Analysed (n= 24)
♦ Excluded from analysis (n= 0)

**TMR group**

**Control group**

Excluded (n= 331)

♦  Not meeting inclusion criteria (n= 331)

♦  Declined to participate (n= 0)

♦  Other reasons (n= 0)

Allocated to intervention (n= 26)

♦ Received allocated intervention (n= 24)

♦ Did not receive allocated intervention (voluntary withdrawal from study) (n= 2)

Allocated to intervention (n= 25)

♦ Received allocated intervention (n= 24)

♦ Did not receive allocated intervention (voluntary withdrawal from study) (n= 1)

Randomized (n= 51)

Assessed for eligibility (n= 382) )

Enrollment

Allocation

Analysis
